# Axes of Prognosis: Identifying Subtypes of COVID-19 Outcomes

**DOI:** 10.1101/2021.03.16.21253371

**Authors:** Emma Whitfield, Claire Coffey, Huayu Zhang, Ting Shi, Xiaodong Wu, Qiang Li, Honghan Wu

**Affiliations:** Health Data Research UK, London, United Kingdom; Institute of Health Informatics, UCL, London, United Kingdom; University of Cambridge, Cambridge, United Kingdom; Usher Institute, University of Edinburgh, United Kingdom; Shanghai East Hospital, Tongji University, Shanghai, China

**Author notes:** These authors contributed equally to this work (Emma Whitfield, Claire Coffey).

## Abstract

COVID-19 is a disease with vast impact, yet much remains unclear about patient outcomes. Most approaches to risk prediction of COVID-19 focus on binary or tertiary severity outcomes, despite the heterogeneity of the disease. In this work, we identify heterogeneous subtypes of COVID-19 outcomes by considering ‘axes’ of prognosis. We propose two innovative clustering approaches − ‘Layered Axes’ and ‘Prognosis Space’ – to apply on patients’ outcome data. We then show how these clusters can help predict a patient’s deterioration pathway on their hospital admission, using random forest classification. We illustrate this methodology on a cohort from Wuhan in early 2020. We discover interesting subgroups of poor prognosis, particularly within respiratory patients, and predict respiratory subgroup membership with high accuracy. This work could assist clinicians in identifying appropriate treatments at patients’ hospital admission. Moreover, our method could be used to explore subtypes of ‘long COVID’ and other diseases with heterogeneous outcomes.

## Introduction

The clinical heterogeneity within COVID-19 patient outcomes has been demonstrated^1,2^, however, a great deal of this heterogeneity remains to be explored. Work has been done to classify patients and predict patient outcomes, much of which has been analysed by Wynants et al.^3^. However, this is often limited to predicting a binary or tertiary severity outcome (death/ICU/none). Our work furthers this by embarking on the exploration of more diverse poor prognosis outcomes.

For a variety of diseases, valuable insights can be gained by using unsupervised clustering to explore prognosis^4,5^. We propose and demonstrate a pipeline for exploring COVID-19 prognosis by using unsupervised clustering methods on patient outcomes to identify subtypes of poor prognosis. The identification of these subtypes facilitates the prediction of patient pathways to nuanced poor prognoses. Our aim is thus to discover and predict a broader range of subtypes of COVID-19 prognosis, using a combination of supervised and unsupervised learning techniques.

## Methods

Our methodology for this task consists of three steps: (1) Feature extraction from multimodal data: exploring, cleaning and manipulating our multimodal dataset in order to extract rich features; (2) Clustering on outcomes: using features collected at endpoints (discharge or death) to cluster patients, comparing outcomes from a variety of clustering approaches using multiple axes of prognosis; (3) Classification at admission: using features at admission, mapping each patient to a cluster found in the previous step to predict their probable deterioration pathways. The pipeline of our work process is shown in Figure 1. We will discuss each step in more detail below. Further details and code are available at https://github.com/knowlab/covid-subtypes.

**Figure 1:**
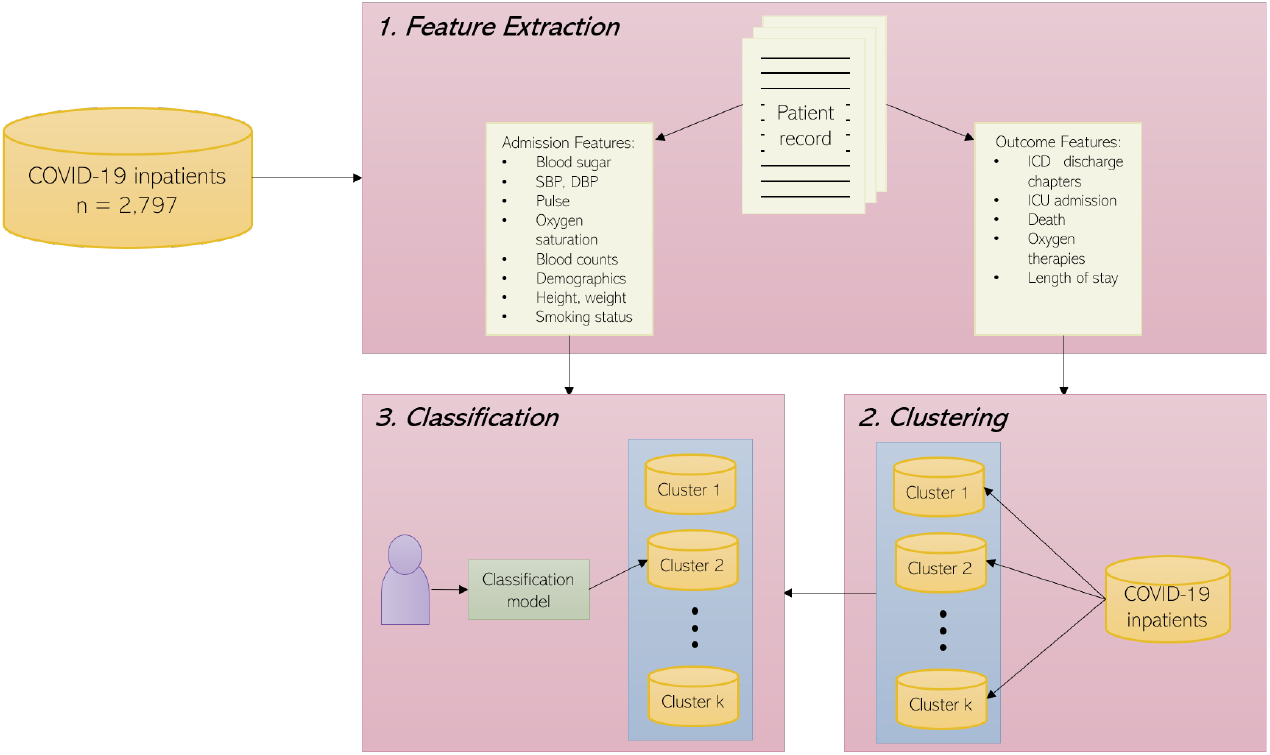
The system architecture to discover and predict COVID-19 subtypes.

### Data

We demonstrate our methodology on a dataset consisting of 2815 health records of COVID-19 inpatients of Wuhan Sixth Hospital and Taikang Tongji Hospital, with admission dates between 4th February 2020 and 30th March 2020. It is worth noting that this cohort was treated in a period with a *treat-all* policy at Wuhan, meaning that admission was routine for all COVID-19 patients. As a consequence, this cohort contains largely non-severe patients and the mortality rate of this cohort is 2.4%.

### Pre-processing

We removed duplicate records, leaving 2797 patients for whom a wide range of features were available, including general patient information, co-morbidities, smoking status (from free-text), symptoms (from free-text), lab test results, ICD-10 admission and discharge codes^6^, and other prognosis features (death, ICU admission, supplementary oxygen, length of stay). Discharge ICD-10 codes were pre-processed and grouped into chapters^7,8^. We removed codes ‘U07.1 - COVID-19’ and ‘Z22 - Carrier of infectious disease’ as, predictably, these were reported for the majority of patients. Discharge conditions were represented as one-hot vectors.

### Data Imputation

For the classification step, we wished to utilise much of the data available on patient admission, including laboratory test results and clinical measurements. However, we did not have readings for each measurement for each patient and so we imputed missing values. Our imputation technique was to randomly choose a value within the normal range for each test result, in order to reduce biasing the results. The values for normal ranges were found from various medical reference sources, the full list of which can be found at https://github.com/knowlab/ covid-subtypes. We chose this, rather than using the average feature value, as we were advised by clinicians working at the hospitals that the absence of values is informative: a test is more likely to be performed if it is suspected the result will be outside the normal range.

### Feature Extraction

To prevent data leakage, we split features into two disjoint subsets: ‘outcome’ features for clustering and ‘admission’ features for classification. Outcome features took binary values, with the exception of length of stay, which was converted to two binary values: 00 for 0-2 weeks, 01 for 2-4 weeks, and 10 for 4 weeks+ (11 is not used). The admission features were taken from measurements in the first three days of hospital admission. Where available, the minumum and maximum readings for days 1 and 3 were used as separate features, otherwise, the average values from each day (1 and 3) were used.

### Discovering COVID-19 Subtypes via Clustering

#### Axes of prognosis model

We wished to cluster on features related to a patient’s outcome in order to explore nuanced subtypes of poor prognosis. To do this, we considered the idea that there are multiple ‘axes’ to prognosis. For example, the two ‘axes’ that make up an individual’s prognosis might be the location of deterioration (defined as clusters of diseases) and the severity of deterioration (quantified/qualified by proxy variables like mortality, types of treatments and length of stay). Here, if a patient had a respiratory discharge code, this would indicate the *location* of deterioration, whilst the use of oxygen therapy would be an indicator of the *severity*. To get a true picture of an individual’s prognosis, we must have a measure across all axes of prognosis - for example, ICD-10 chapters to indicate area, and features such as ICU admission, death, oxygen therapies, and length of stay to measure severity. We will use different clustering approaches to leverage the idea of ‘axes of prognosis’.

The following example illustrates the potential value of this approach. Consider three patients who are admitted with COVID-19: patient A has no significant symptoms and is discharged after 5 days; patient B arrives with no significant symptoms, but is noted to be vitamin D deficient during their stay, recorded as ‘E55.9’, they are discharged after 7 days; patient C has already deteriorated significantly upon arrival and dies 1 day later. Clustering algorithms typically use distance measures to compare data points. If we consider these three patients in the space ‘Has Nutritional Code’ *×* Death = {0, 1}^2^ then, for most distance measures *d, d*(patient A, patient B) = *d*(patient A, patient C). Thus, clustering algorithms struggle to note the clearly significant distinction between outcomes. On the other hand, were we to consider the axes ‘location of deterioration’ and ‘severity of deterioration’ individually, or to transform the features into some space where distance can be ‘sensibly’ measured, a clustering algorithm could note the difference in severity of deterioration between patient C and patients A and B.

We propose the use of two novel methods using axes of prognosis: *Layered Axes* and *Prognosis Space* with the goal of exploring nuanced subtypes of poor prognosis. These are summarised below and in Figure 2.

**Figure 2:**
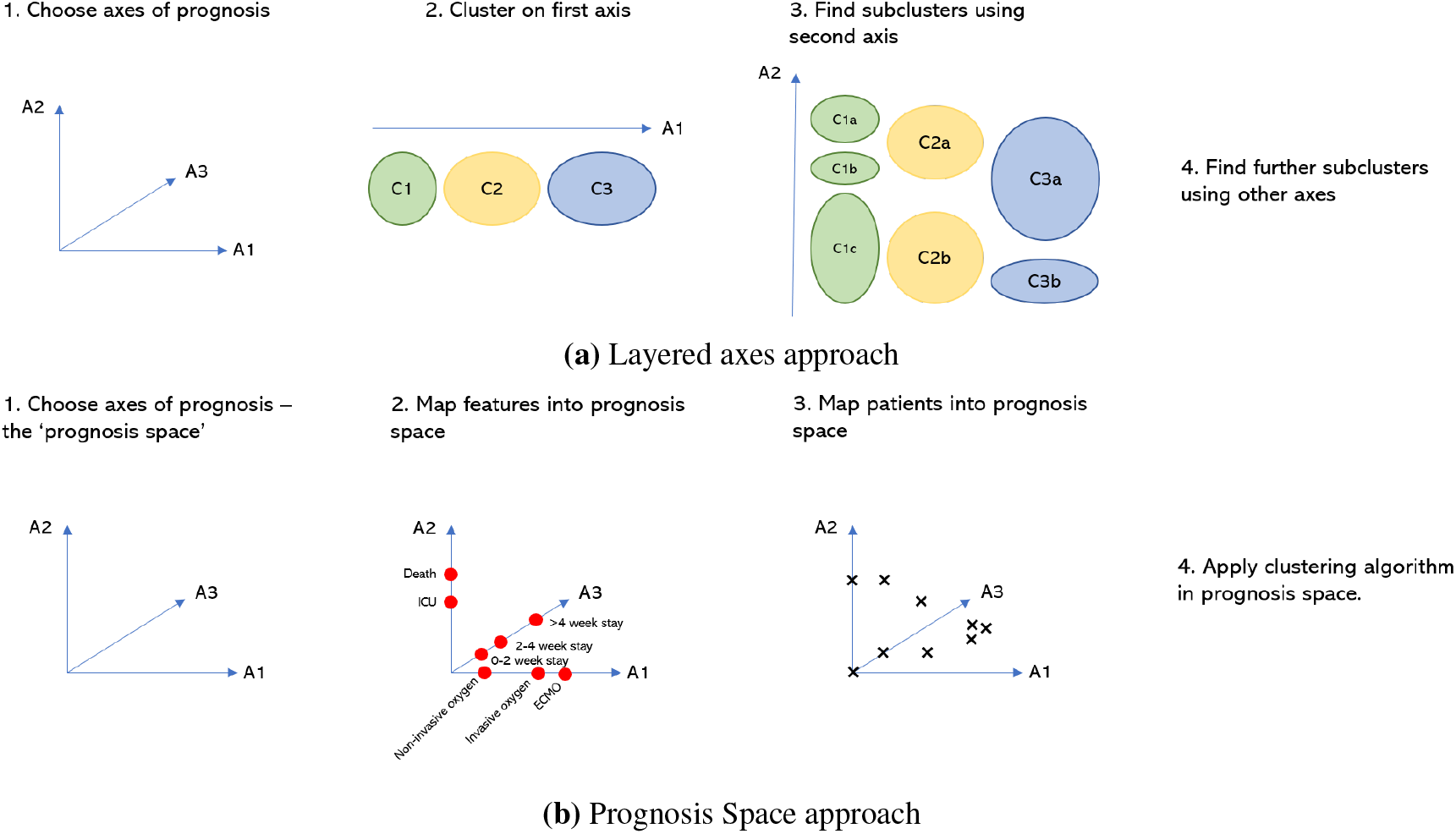
Two approaches to clustering using axes of prognosis.

##### Layered Axes

1. Choose axes of prognosis (*e*.*g. location of deterioration, severity of deterioration, duration of illness*) and assign each outcome feature to the axis of which it is most indicative (*for example, ICD-10 chapters are indicative of the location of deterioration*).
2. Choose one axis and, using only the features assigned to it, apply a standard clustering method to produce clusters. This produces clusters that clearly describe that axis.
3. Discard the features already used and choose a new axis. For each cluster already found, use the features assigned to the new axis and a standard clustering method to produce subclusters. These subclusters now clearly describe both axes.
4. The subclusters become the clusters and repeat step 3 until all axes have been considered.

##### Prognosis Space

1. Choose axes of prognosis and assign each outcome feature to the axis of which it is most indicative.
2. Define a ‘space transformation’ function to map features onto the axes - creating an interpretable Prognosis Space with smaller dimensions. This allows domain knowledge to be incorporated into the dimension reduction.
3. Use this function to map patients into the Prognosis Space.
4. Apply a clustering algorithm in the Prognosis Space. As we are no longer in a binary feature space, common distance measures, such as the Euclidean distance, have more meaning - this allows us to apply a wider range of clustering algorithms, such as DBSCAN^9^.

#### Implementation

Our dataset consists of 2797 patients, for whom we have 16 binary features describing their outcome. As a baseline, we applied a standard clustering method to all features. Then, we applied both of our new methods to the Wuhan dataset and compared the clusters produced. For the Layered Axes method, we chose our axes of prognosis to be location and severity of deterioration. We used ICD-10 chapter codes to indicate location of deterioration and ICU admission, death, use of noninvasive/invasive oxygen therapy and ECMO, and length of stay to indicate severity of deterioration. We clustered first on the location and then found subclusters using severity.

For the Prognosis Space method, we chose our axes of prognosis to be location of deterioration, need for oxygen therapy, severity of deterioration and duration of illness. As above, ICD-10 codes were used to indicate location; length of stay indicated duration of illness, ICU admission and death indicated severity of deterioration, and use of oxygen therapy and ECMO indicated need for oxygen therapy. We created a ‘space transformation’ function, *f* : {0, 1} ^16^ →ℝ^4^, that placed more weight on the severity and need for oxygen therapy axes. The function has the basic form *f* (**x**) = (*f*_1_(**x**_location_), *f*_2_(**x**_OXY_), *f*_3_(**x**_severity_), *f*_4_(**x**_duration_)), where, for example, **x**_location_ refers specifically to the features of **x** that we used to indicate location. More details of the dimension reduction function used are given at https://github.com/knowlab/covid-subtypes.

As all our outcome features were binary and we wished to demonstrate the potential utility of this method in a clinical setting, we used the K-Modes clustering algorithm^10^ (implemented using the Python package kmodes^11^) for our baseline and Layered Axes approach. This forms clusters based around a ‘centre’ - specifically the mode of the cluster - and uses the Hamming distance to compare points. The cluster centres are actual data points, which can then be used as a description for each cluster produced. The algorithm was run with several different numbers of clusters and the best result for each approach was chosen to reflect the heterogeneity in outcomes. Other clustering methods - such as K-Means clustering and DBSCAN - were also tested, however these algorithms are not optimised for the binary features used and so were less interpretable in our context.

Our Prognosis Space approach maps the binary features into ℝ^4^ - this means it now makes sense to use the Euclidean distance to compare points, and a wider range of clustering algorithms become suitable. We demonstrated our Prognosis Space result using DBSCAN as it is more capable of capturing clusters of different shapes. Hyperparameters were optimised using the elbow method.

#### Cluster Analysis

As this task is unsupervised, we had no knowledge of what ‘good’ clusters look like. Therefore, we sought clusters with a clear clinical interpretation, showing distinctions between different prognoses. Our cohort contained a large number of patients with mild symptoms: discounting length of stay, 1744 patients had 0s for all features. As such, for this cohort we expected at least one cluster with over 1000 patients, with others significantly smaller in size. To determine the clinical interest of a set of clusters, we examined heatmaps of feature prevalence for all the outcome features, alongside demographic feature prevalence.

### Predicting COVID-19 Subtype at Admission via Classification

For prediction, we used the clusters discovered as labels to generate a supervised classification problem. This not only provides information about potential patient deterioration pathways, which can be used to assist clinical decision making (e.g. how best to treat a patient), but also provides a way to validate the clinical meaning of the clusters found.

#### Poor Prognosis Subtypes

Since we were most interested in predicting the subtypes of poor prognosis, we first combined the clusters into two categories: ‘non-severe’ and ‘severe’ and performed a binary severity classification. These categories were decided using our understanding of the clinical meaning of the clusters. Then, in order to examine subtypes of prognosis, we focused only on the interesting patients. We disregarded individuals with no recorded discharge codes (since they had no prognosis subtypes), and performed multiclass classification on the remaining patients to predict their deterioration pathway. We tested decision trees, logistic regression, and random forest classifiers - all transparent models traditionally used in clinical decision making - since explainability is important. We will present the results found using random forests as these provided the best performance.

#### Classification Models

For our system to be clinically useful, it is imperative the models are interpretable to clinicians. This motivated our choice of a set of ‘transparent models’ including decision trees, logistic regression and random forests^12^. We only present results on random forests in this paper as it performed the best. We used scikit-learn’s^13^ implementation. Random forests utilise many decision trees as an ensemble, which reduces potential issues with overfitting found in decision trees; they can provide better results than single decision trees since the variance between predictions is captured in the overall model. The space used and runtime is greater than that of a single decision tree, but this was not an issue for us due to the small dataset. The trees can be plotted, clearly showing decisions made by the classifier, including the importance of each feature used to aid in these decisions.

#### Classification Details

We implemented grid search to choose the optimal hyperparameters for each classifier, and we used scikit-learn’s^13^ stratified K-fold cross-validation to validate our models. The use of cross-validation was important due to the small cohort size: we wanted to make best use of the data by training and testing the model on multiple combinations of our data. The stratified variant of K-fold preserves the underlying distribution of the data, which we used due to the class imbalance observed. The value of *k* was set to 3; this was kept small to ensure there were enough samples in the test data for the least numerous classes in each fold.

#### Addressing Data Imbalance

A limitation for the success of classification is the class imbalance that arises from many patients experiencing only mild symptoms. Since we are using real-world data, the imbalance of classes is representative of a real Chinese hospital cohort. However, to optimise the generalisability of our classifiers and increase performance, we used the following techniques on our training data:

##### Downsampling

We downsampled individuals in the most numerous classes by randomly removing a proportion of these from the training data for each fold. Overall predictions are improved, but reducing the size of the training data means nuances are less likely to be captured. This was used only for the binary severity classification.

##### Upsampling

We upsampled individuals in the least numerous classes in the training data in each fold by randomly sampling with replacement. Overall predictions are improved, however upsampling does not capture the variety of patients, potentially causing overfitting and not improving predictions for those deviating from the training set.

## Results

### Clustering

For the baseline and Layered Axes approaches, we used the K-Modes cluster centre descriptions for each cluster. For the Prognosis Space approach, we derived similar descriptions using the heatmaps produced. The descriptions of all the clusters found are given in Table 1, and heatmaps of the prevalence of binary features in each cluster are shown in Figure 3.

**Table 1:** Clusters found using the three approaches - the standard baseline clustering approach, the Layered Axes approach and the Prognosis Space approach. Interesting subtypes are highlighted in bold text.

| Approach | Clustering Method | Cluster Label | Size | Description |
| --- | --- | --- | --- | --- |
| **Baseline** | **K-Modes** | 0 | 73 | Digestive, los 2-4 weeks |
|  |  | 1 | 1374 | Non-severe |
|  |  | 2 | 97 | Respiratory, ICU, death, oxygen therapy |
|  |  | 3 | 1018 | Los 2-4 weeks |
|  |  | 4 | 173 | Circulatory |
|  |  | 5 | 62 | Digestive |
| **Layered Axes** | **K-Modes** | 0a | 150 | Nutritional, non-severe |
|  |  | 0b | 22 | **Nutritional, ICU, Death, Oxygen therapy** |
|  |  | 0c | 113 | Nutritional, los 2-4 weeks |
|  |  | 1a | 45 | **Respiratory, ICU, oxygen therapy** |
|  |  | 1b | 131 | **Respiratory, non-severe** |
|  |  | 2a | 90 | Circulatory, los 2-4 weeks |
|  |  | 2b | 120 | Circulatory, non-severe |
|  |  | 3a | 845 | Non-severe, los 2-4 weeks |
|  |  | 3b | 1281 | Non-severe |
| **Prognosis Space** | **DBSCAN** | -1 | 37 | Noise |
|  |  | 0 | 914 | Non-severe, los 2+ weeks |
|  |  | 1 | 1114 | Non-severe, los <2 weeks |
|  |  | 2 | 112 | **Oxygen therapy, los 2+ weeks** |
|  |  | 3 | 255 | Circulatory, los <2 weeks |
|  |  | 4 | 232 | Circulatory, los 2+ weeks |
|  |  | 5 | 43 | **Respiratory, ICU, no death** |
|  |  | 6 | 44 | Oxygen therapy, los <2 weeks |
|  |  | 7 | 28 | **Respiratory, death in <2 weeks** |
|  |  | 8 | 18 | **Many side effects, death in 2+ weeks** |

**Figure 3:**
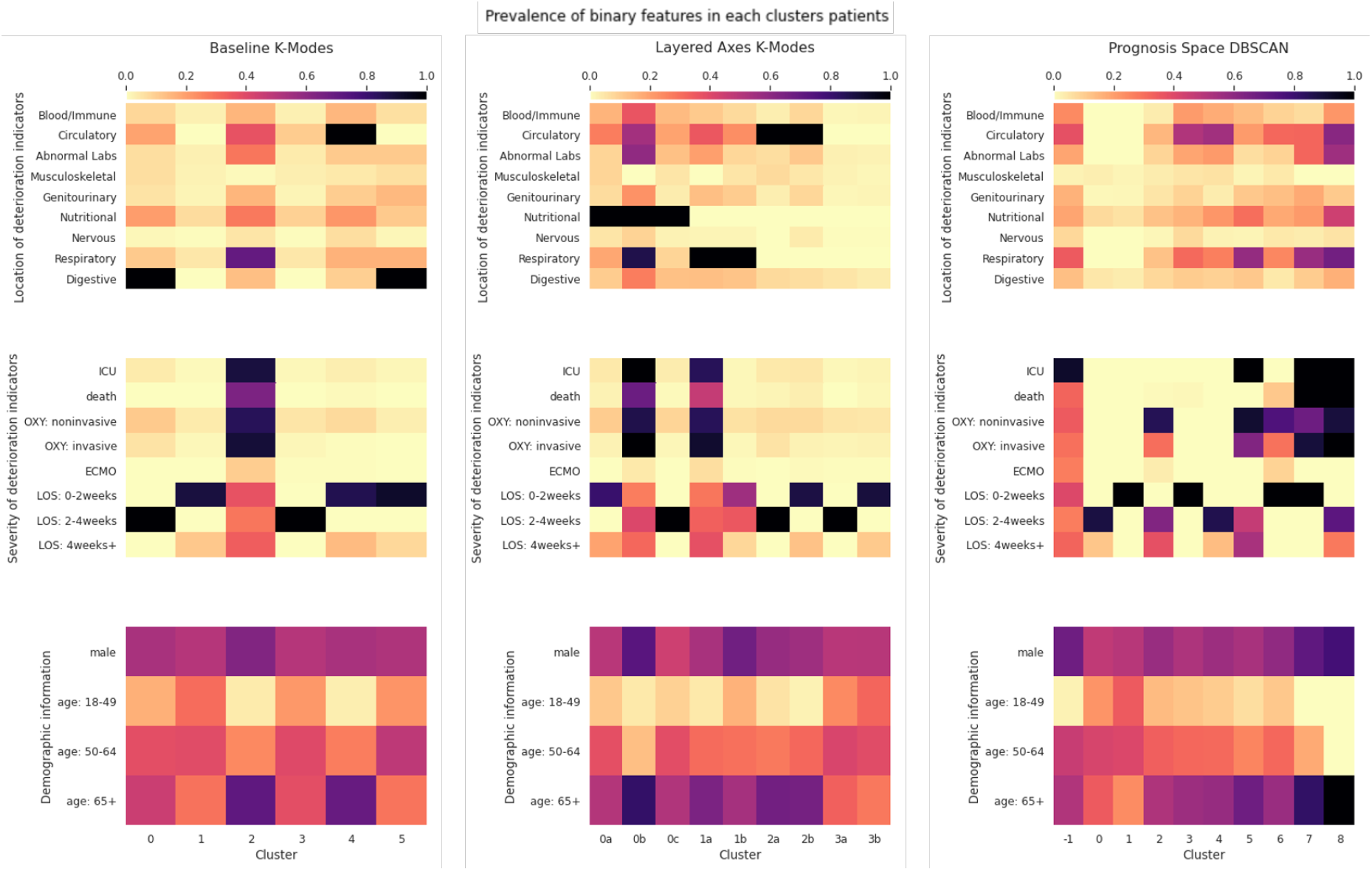
Heatmaps showing prevalence of binary features within each cluster found by each approach. OXY refers to use of oxygen therapy and LOS refers to the length of stay. Note that the features shown in the third row, Demographic information, were not used for clustering - they are shown only to aid interpretation of the clusters. For the Layered Axes K-Modes clusters, labels are in the form number-letter where the number indicates the clusters found from the first axis, and the letter the subclusters found within that cluster using features from the second axis. Note also that for the Prognosis Space approach, DBSCAN has been used and −1 indicated points described as ‘noise’.

#### Baseline Approach

Most severe patients were put in cluster 2, meaning it failed to find subtypes of severe cases. There are two non-severe subtypes with no adverse prognoses: cluster 3 with 2-4 weeks of stay and cluster 1 with mixed stays, most within 2 weeks and a few 4+ weeks.

#### Layered Axes

This method discovered several clinically sensible subtypes within groups with similar severity and background conditions. There are two clear subtypes of severe COVID patients: cluster 0b with high proportion of co-morbidities, particularly a combination of respiratory and nutritional; cluster 1a with mostly respiratory that were complicated with circulatory; the former was generally more severe. There are two subtypes of respiratory patients: cluster 1a - the severe group and cluster 1b - largely non-severe (40% discharged within 2 weeks). This is particularly interesting as respiratory conditions are widely recognised as a high risk-factor of severe COVID-19 cases despite the age group. There are also two subtypes of non-severe patients: cluster 3a with 2-4 weeks of hospital stay and cluster 3b with around 80% discharged in 2 weeks with no recorded complications.

#### Prognosis Space

There are three very interesting subtypes of severe patients: cluster 5 - all stayed in ICU but all recovered; cluster 7 - quick deterioration to death within 2 weeks; cluster 8 - others all died after *>*2 weeks. For non-severe patients: cluster 1 patients have no underlying conditions and are discharged within 2 weeks; cluster 3 have mostly circulatory conditions (also many other conditions) and a speedy recovery (*<*2 weeks of stay); cluster 0 - others, with longer admission. There is another distinct finding - cluster 2 were those patients mostly on oxygen therapy but who recovered and never stayed in ICU.

### Classification

We split the classification procedure into two classification tasks:

1. Predicting binary severity (severe/non-severe) using baseline K-Modes clusters.
2. Predicting poor prognosis subtypes of ‘interesting’ patients. This contained classification sub-problems, based on the labels generated by Baseline K-Modes, Layered Axes K-Modes and Prognosis Space DBSCAN.

For each task, we classified using random forests. We implemented grid search in order to find the optimal hyper-parameters. These, along with additional experimental configurations can be found at our online resource: https://github.com/knowlab/covid-subtypes. Below is a description of the experiments and their results.

#### Binary Severity Classification

To classify severity, we grouped clusters into ‘severe’ and ‘non-severe’ categories. A summary of the clusters is presented in Table 2, along with their death and ICU statistics, which show a clear difference in the attributes of the two severity levels. Our classification results are shown in Table 3. We present these for only the baseline K-Modes clustering, since the binary classification is not the main focus of this work; we give precedence to our nuanced prognosis subtypes prediction. We achieved an *F*_1_ score of **0**.**722** for the binary classification to identify the most severe patients (experiencing ICU or death) vs non-severe (all other patients).

**Table 2:** Baseline K-Modes clusters for binary classification.

| Severity | Clusters | Size | Death % | ICU % |
| --- | --- | --- | --- | --- |
| Non-severe | 0, 1, 3, 4, 5 | 2700 | 0.259 | 1.370 |
| Severe | 2 | 97 | 63.918 | 88.660 |

**Table 3:** Subtype classification results (macro average of multi-label results)

| Approach | Clustering method | Acc. | Recall | Precision | $F_1$ |
| --- | --- | --- | --- | --- | --- |
| Baseline (binary) | K-Modes | 0.969 | 0.686 | 0.781 | 0.722 |
| Baseline (multiclass) | K-Modes | 0.573 | 0.495 | 0.518 | 0.491 |
| Layered Axes (all) | K-Modes | 0.657 | 0.626 | 0.637 | 0.627 |
| Layered Axes (severe) | K-Modes | 0.627 | 0.547 | 0.547 | 0.542 |
| Layered Axes (respiratory) | K-Modes | 0.886 | 0.829 | 0.866 | 0.844 |
| Prognosis Space (all) | DBSCAN | 0.428 | 0.316 | 0.356 | 0.320 |
| Prognosis Space (severe) | DBSCAN | 0.651 | 0.581 | 0.665 | 0.579 |

#### Poor Prognosis Subtypes Classification

Since identifying prognosis subtypes was a main goal of this work, we focused on classifying the ‘interesting’ patients, disregarding patients without discharge codes or poor prognosis events.

##### Layered Axes

The results for the Layered Axes clusters are shown in Fig. 4. These display an improved overall accuracy compared to baseline K-Modes, with an *F*_1_ score of 0.638 on average. We built separate classifiers to predict the second-layer subcluster membership (within each top-layer cluster of digestive, respiratory, and circulatory clusters). The *F*_1_ score for the classification of respiratory patients’ subtypes is highest at **0**.**844**. In figure 5, the confusion matrix for the classification of the most severe patients is shown; the sub cluster sizes are very limited which impacts the robustness of these results.

**Figure 4:**
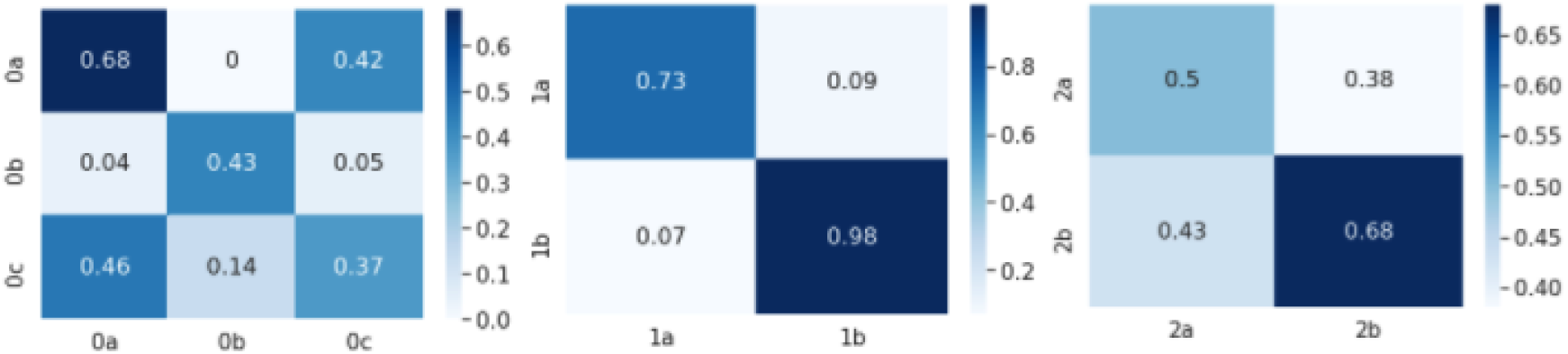
Example confusion matrices heatmaps for Layered Axes K-Modes classification subclusters. Left: Nutritional; Middle: Respiratory; Right: Circulatory. x-axis: predicted label; y-axis: true label

**Figure 5:**
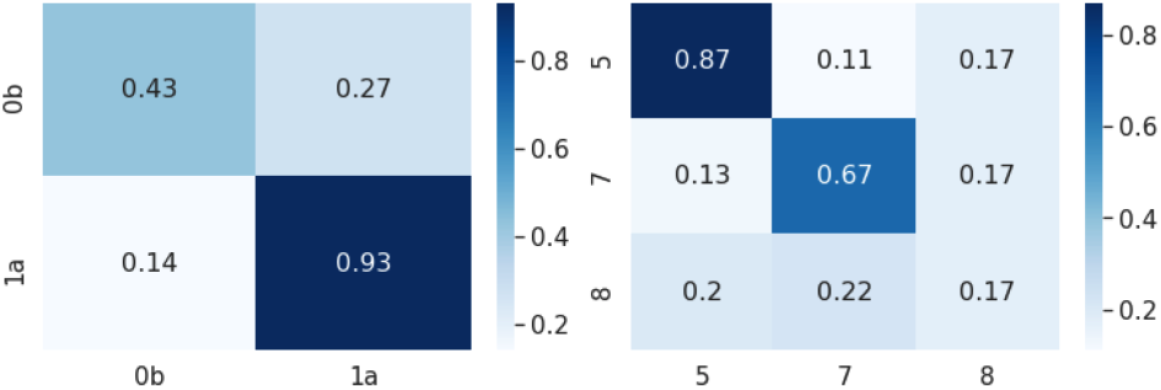
Example confusion matrices for classification of severe patients. Left: Layered Axes K-Modes; Right: Prognosis Space DBSCAN. x-axis: predicted label; y-axis: true label

##### Prognosis Space

We present the results for the classification of severe patients’ subgroups using the Prognosis Space approach in Figure 5. Subtypes 5 and 7 are classified with higher accuracy than 8, although these are the most severe patients. Here, the cluster sizes are very limited which impacts the robustness of these results.

## Discussion

Our approach is able to identify many subtypes of severe COVID-19 cases described with rich clinical features. We believe this is the first work that reveals the heterogeneity of COVID-19 with such detail. Many of these subtypes (such as clusters 1a and 1b for Respiratory patients) were surfaced for the first time, conveying novel insights that lead to better understanding of the disease and could potentially guide us to treat patients with more personalised approaches. It is clear that the axes of prognosis approach plays a vital role in achieving this.

Comparing to baseline K-Modes, using the Layered Axes approach can discover more nuanced clusters by grouping on one axis at a time and thus selecting a feature of significance from each axis. For example, at baseline, respiratory side effects are only considered in the context of very severe deterioration. In contrast, clusters 1a and 1b capture different outcomes for respiratory patients. It is interesting to note that, despite the very different outcomes of 1a and 1b, the proportion of over 65s is similar in both clusters. We might have expected that older patients were more likely to deteriorate significantly. Prognosis Space is also able to reveal more heterogeneity in outcomes. Clusters 5-8 seem particularly noteworthy for their similar yet distinct characteristics. In particular, clusters 7 and 8 both consist of patients who are admitted to ICU and die, but with quite distinct deteriorating speed.

For classification, in the binary problem, the *F*_1_ score achieved is 0.722, showing that we can identify the most severe patients with good accuracy. In Layered Axes, the classification results suggest that these nuanced multi-axis clusters not only make more clinical sense, but they also provide improved classification results in comparison to the baseline. In particular, the excellent classification results for the respiratory subclusters, 1a and 1b, provide a strong example use case. Here, we can predict accurately whether a respiratory patient is likely to deteriorate seriously leading to ICU and death (1a membership) or experience no severe prognosis (1b membership). This is especially interesting since there is no significant difference in the age of the patients between these two subclusters (Fig. 3). For Prognosis Space, results are promising, but lacking in robustness due to the larger number of clusters and therefore the smaller number of samples in each cluster. It is interesting that our classifier can distinguish between clusters 5 and 7 relatively easily (Fig. 5) but struggles with cluster 8. Cluster 8 contains patients who deteriorate fatally, but not rapidly. Our classifier likely struggles with these individuals since they take a longer time to deteriorate (in contrast to cluster 7) so their readings upon admission - which we use as classification features - may not be seriously irregular.

### Strengths

We have demonstrated that considering axes of prognosis allows for discovery of more nuanced subtypes of poor prognosis. In particular, our Layered Axes approach allows us to interpret every cluster found along each axis of prognosis being considered and is easily interpretable, especially when only a small number of axes are being used. On the other hand, our Prognosis Space approach allows us to incorporate domain knowledge and combine related binary features in a meaningful way. The Prognosis Space approach is comparable to other dimension reduction-clustering approaches such as Prinicipal Component Analysis (PCA). We believe that compared with methods such as PCA, especially in the context of a binary feature space, our approach is both more interpretable and allows for the incorporation of targeted domain knowledge. Both of our approaches provide a more interpretable and clinically meaningful output than many baseline clustering approaches.

### Limitations

We highlight the fact that the large majority of our cohort did not suffer from any significant side-effects from COVID-19. In fact, the clusters contain over 2300 patients who can be deemed to be ‘non-severe’ (clusters 1 and 3 in baseline K-Modes). This leaves only around 500 patients from whom we can derive more ‘interesting’ clusters and build classification models, leading to classifiers built from very small samples. Upsampling the smaller classes when training our models cannot capture diversity in test samples, so does not appear to be a good solution here. Therefore, our approach, especially when using the clusters found across multiple axes, must be tested on larger datasets.

Additionally, although we have extrapolated meaning from the clusters found, we cannot truly know which clustering is ‘best’, or even, ‘good’ since they are found in an unsupervised setting. If clustering is not optimal, our classification will likely also be worse - but this is a hard problem to overcome! A larger and more diverse dataset may also help with our confidence in clustering ability and predictions made.

### Future Work

This work provides a demonstration of our methodology for exploring heterogeneous disease prognosis. Therefore, potential future work is vast. Predominantly, these techniques need to be tested on a larger dataset, particularly with more patients with severe outcomes. This will likely improve the accuracy of clustering on severe patients, and the ability of classifiers to predict them. The methodology behind the Prognosis Space technique in particular should be applied to a larger dataset to properly explore its capabilities. Furthermore, a large dataset would allow for deep methods such as neural networks to be tested - these have been shown to perform very well on unsupervised learning problems with large datasets. Whilst we would not want to use these for our main predictions due to lack of interpretability, they could provide a useful comparison.

Secondly, our model may not be generalisable to cohorts from other countries, due to the different policies on hospital admission. However, we believe this approach could remain useful and provide interesting insights. Testing on different cohorts would enable exploration of the generalisability of the work.

Further to this, our methodology needs to be tested and explored more rigorously. In different contexts, there are a vast number of clustering algorithms that could be used within our method - especially when continuous features are used. Moreover, the methodology could be refined and optimized for different contexts and diseases.

In some cases, classifiers could confidently predict patients who would die in a short time frame, yet struggled to identify those who would die after a longer period of deterioration, even if the outcomes were similar. Another aim for future work could be to establish whether using later time series points for these patients would allow correct classification. We could also explore whether this would have potential applications for predicting long COVID. In particular, if a patient took a certain non-fatal prognosis pathway, this could be linked to the development of specific symptoms of long COVID later on.

## Ethics Declaration

This study was approved by the Research Ethics Committee of Shanghai Dongfang Hospital.

## Conclusion

We have proposed a novel *axes of prognosis* model and demonstrated how it could be used to identify diverse COVID-19 prognosis that accounts for the wide heterogeneity of the disease via a combination of clustering and classification. This methodology is not only suitable for predicting prognosis, but has a wide range of potential applications, for example, risk prediction and helping clinicians take preventative measures earlier on. This methodology could also be easily applied to a range of diseases with heterogeneous prognoses, including studying subtypes of long COVID.

## Data Availability

Data pre-processing, machine learning details and all relevant scripts are available at https://github.com/knowlab/covid-subtypes. Details of the cohort are described at https://covid.datahelps.life/. The patient data used in the study will not be available due to inability to fully anonymize in line with ethical requirements. Applications for research access should be sent to TS.

https://covid.datahelps.life/

